# Efficacy of Acupuncture on Lower Limb Motor Dysfunction Following Stroke: A Systematic Review and Meta-Analysis of Randomized Controlled Trials

**DOI:** 10.1101/2024.10.18.24315720

**Authors:** Peng Chen, Xing Jin, Debiao Yu, Xiaotin Chen, Yaoyu Lin, Fuchun Wu, Bin Shao

## Abstract

**Background:** Acupuncture is widely used for Lower Limb Motor Dysfunction Following Stroke (LLMD) in China but its effect is unclear. We aim to evaluate the effect of acupuncture for LLMD.

**Methods:** We searched eight databases, including PubMed, Embase, Web of Science, Cochrane Library, China National Knowledge Infrastructure, Wanfang, VIP, and CBM, up to December 2023. Randomized controlled trials on acupuncture therapy for LLMD after stroke were enrolled in this study. Outcome measures comprised motor function, balance function, walking ability, and daily living activities. Two researchers conducted independent literature screening, data extraction, and quality assessment in accordance with Cochrane Collaboration network’s standards. Review Manager 5.3 and Stata 17.0 were used in data analysis. Results were presented as mean difference (MD) or standardized mean difference (SMD) with 95% confidence interval (95% CI).

**Results:** Twelve studies involving 1318 patients, most of which showed low or unclear risk of bias, were included in this review. Meta-analysis results indicate that compared with conventional treatment, acupuncture intervention can improve scores in Fugl–Meyer Assessment for lower scale (SMD=0.41, 95% CI (−0.92, –0.04), Z=2.16, P =.03), Berg Balance Scale (SMD=-0.86, 95%CI (−1.65, –0.07), Z=2.14, P=.03), Functional Ambulation Category scale (SMD=-0.74, 95%CI (−2.33, 0.84), Z=0.92, P=.36), and Modified Barthel Index Scale (SMD=0.27, 95%CI (−0.30, 0.84), Z=0.94, P=.35).

**Conclusion:** The results of this study suggest that acupuncture combined rehabilitation training may be more effective than pure treatment in improving LLMD, balance function, walking ability, and daily living activities after stroke. Despite limitations due to the low quality of the included studies and methodological constraints, acupuncture combined rehabilitation training may serve as an effective approach for the treatment of LLMD poststroke.

## 1. Introduction

A cerebral incident, often referred to as a cerebrovascular accident (CVA), transpires when an obstruction impedes cerebral blood flow or during cerebral hemorrhage due to the rupture of a blood vessel within the brain^1^. Approximately 12.2 million new cases of stroke are recorded each year, with over 80 million stroke survivors globally^2^. Moreover, the incidence of stroke, which severely impacts individual health, is increasingly affecting younger populations^3^. Hemiparesis is a typical sequela among survivors, with over 50% experiencing substantial lower limb motor dysfunction (LLMD)^4^. This condition greatly compromises a patient’s ability to walk and maintain posture stability, which considerably affect their quality of daily life^5^. Impaired lower limb motor function also hampers the rehabilitation process, diminishes individual autonomy, and restricts the patients’ ability to engage in social roles, which further exacerbate their physical and psychological burden^6,7^. Various treatments, including pharmacotherapy, physical therapy, rehabilitation training, assistive devices, and supportive equipment, have been developed to address poststroke motor deficits. However, these interventions pose different side effects and may decrease patience and confidence of patients due to prolonged rehabilitation training^8-10^.

Acupuncture, which originating in China, is a traditional treatment method based on the theory of meridians in traditional Chinese medicine^11^. Acupuncture therapy has been widely used in the treatment of movement disorders secondary to diseases, such as stroke, brainstem injury, Parkinson’s disease, spinal cord injury, multiple sclerosis, and peripheral nerve and muscle injuries^12,13^. Animal studies have shown the improved motor function of patients with various diseases, including poststroke motor dysfunction, due to acupuncture, which regulates the nervous system function, promotes blood circulation, and relieves muscle spasms with few side effects and risks^14-16^.

Clinical studies have demonstrated the capability of acupuncture to improve walking ability and lower-limb balance, strengthen lower-limb muscles, and enhance the gait of stroke patients^17,18^. However, other research suggest the nonsignificant recovery effect of acupuncture on poststroke LLMD^19^. This controversy requires further validation. Therefore, a meta-analysis must be conducted to evaluate the therapeutic effects of acupuncture on LLMD after stroke and assess its effect on motor function, balance function, walking ability, and daily living activities to provide evidence-based medicine for clinical applications of acupuncture in the treatment of LLMD after stroke.

## 2. Materials and Methods

This study, being a systematic review and meta-analysis, primarily involves the re-examination of data from previously published research rather than direct engagement with patients or access to personal health information. As such, under international guidelines for research ethics, this type of investigation does not require fresh ethical approval or patient consent.

### 2.1. Study registration

This protocol has been registered in the International Prospective Register of Systematic Reviews (PROSPERO) under trial registration number CRD42023487617.

### 2.2. Search strategy

The search strategy was implemented in accordance with the Cochrane Handbook guidelines (5.1.0). From inception to December 31, 2023, we searched the following electronic databases: Embase, PubMed, Cochrane Library, Web of Science, China National Knowledge Infrastructure, Wan Fang, China Science Journal Database (VIP), and Chinese Biomedical Literature Database. In addition, ongoing unpublished trial data were searched from clinical trial registries, such as the WHO International Clinical Trials Registry Platform and the Chinese Clinical Trial Registry. The corresponding authors were contacted in case of incomplete data. The search strategy was adjusted based on the combinations of subject headings and free terms used in each database. For specific retrieval strategies, take PubMed as an example, as shown in Table 1.

**Table 1.**
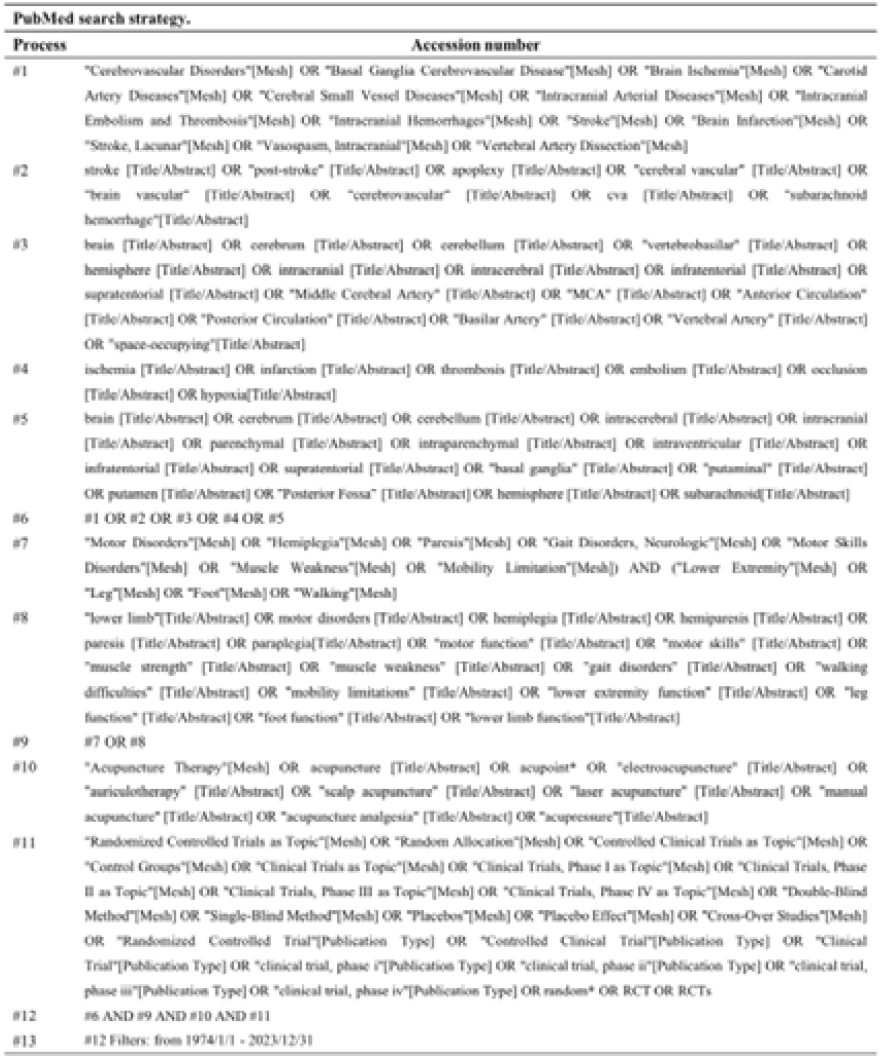
PubMed search strategy.

[Insert keywords for other databases (Appendix 1)].

### 2.3. Inclusion criteria

#### Study Type

Randomized controlled trials (RCTs) investigating the effects of acupuncture on poststroke lower limb motor function and limited to publications in Chinese and English.

#### Study Participants

Patients aged 18–80 years with confirmed stroke via cranial brain computed tomography or magnetic resonance imaging, exhibiting LLMD post-stroke, with clear consciousness, and capable of cooperation during examinations and treatment.

#### Interventions

The control group received routine training (rehabilitation, sensory motor, drug therapy + rehabilitation, rehabilitation, and functional training); the observation group underwent acupuncture treatment.

#### Outcome Measures

Motor function, balance function, walking ability, and daily living activities assessed through the Fugl–Meyer Assessment for Lower Scale (FMA-L)^20^, Berg Balance Scale (BBS)^21^, Functional Ambulation Category scale, and Modified Barthel Index scale, respectively.

### 2.4. Exclusion Criteria

Duplicate publications; non-RCTs including animal experiments, literature reviews, systematic reviews, meta-analyses, case reports, expert summaries, operational research, parameter studies, prospective or retrospective clinical observations, comments, letters, conference abstracts, etc.; useful data cannot be obtained by contacting authors; swallowing difficulties due to other reasons, such as brainstem injury, Parkinson’s disease, spinal cord injury, multiple sclerosis, peripheral nerve and muscle injuries, etc.; the observation group receiving combined treatment methods different from that provided to the control group apart from acupuncture.

### 2.5. Literature selection

Literature search and screening were conducted by two researchers independently. Initially, all literatures retrieved from the preliminary search were imported into EndNote (Clarivate Analytics 21.0), and duplicates were removed using software functions; highly irrelevant literatures were excluded beyond reading titles, abstracts, and keywords. Subsequently, further screening was achieved with the two researchers independently reading the full text of potentially included literature. Those that failed to meet the criteria were deleted, and reasons for study exclusion were recorded. Moreover, a third party was assigned to conduct a reassessment and make a decision when disagreements occurred between the two researchers. The study selection process was described using the Preferred Reporting Items for Systematic Reviews and Meta-Analyses flow diagram the.

### 2.6. Data extraction

The reviewers also independently performed data extraction from the studies. After comparison of their results and verification with the original articles, the accuracy and completeness of each data point were confirmed. Extracted data mainly included author, publication year, sample size, patient age, interventions, treatment process, and outcome measures. The corresponding author was contacted for information that needed supplementation.

### 2.7. Assessment of risk of bias (ROB)

The two independent researchers assessed the risk of bias using the methods endorsed by The Cochrane Collaboration, which included the following domains: (a) randomization process; (b) deviations from intended interventions; (c) missing outcome data; (d) measurement of the outcome; (e) selection of the reported result. Any disagreements were resolved by discussion.

### 2.8. Statistical methods

Statistical analysis was conducted using Review Manager 5.3 software (Cochrane, England). Odds ratio (OR) and 95% confidence interval (CI) were used as combined effect size indicators for categorical variables, and standardized mean difference (SMDs) were applied for continuous variables. SMD and 95% CI were used as indicators of the combined effect size. In case of missing data, studies reporting their outcomes were included in the statistical analysis. In case of incompletely reported required change amount, the reviewers consulted the Cochrane Handbook to perform manual calculations of the mean and standard deviation based on the baseline and outcome data reported. I^2^ and ChI^2^ tests were conducted to determine the heterogeneity of included studies. For studies with zero heterogeneity or low heterogeneity (I^2^ < 50%, P > 0.1), a fixed-effects model was used for analysis. In cases of high heterogeneity (I^2^ ≥ 50%, P < 0.1), a random-effects model (REM) was used for the analysis. In cases of significant heterogeneity, efforts were exerted to identify potential sources and consider subgroup or sensitivity analyses. Descriptive analysis was conducted when data could not be synthesized. Funnel plot analysis and Egger test (performed in Stata 17.0 software) were used to assess the risk of publication bias with respect to outcome measures in ≥10 included studies. If the distribution in the funnel plot was approximately symmetric and the P value of the Egger test was >0.05, it indicated no significant bias and the conclusion was reliable.

## 3 Results

### 3.1 Study selection

The initial search strategy yielded 2718 records for consideration in this work. The initial screening process identified and excluded 1341 duplicates, unqualified items flagged by automated tools, and other reasons. Among the remaining 1377 articles, in accordance with the inclusion criteria, guidelines, animal experiments, reviews, data mining, operational studies, correlational studies, non-acupuncture intervention studies, and non-RCT papers were deleted, and 12 RCTs^22-33^ were finally included. Figure 1 depicts the process and results of literature screening.

### 3.2 Study characteristics

This meta-analysis included 12 trials involving 1318 patients. All papers were published from 2011 to 2023, described as RCTs, and published in Chinese or English. The studies had sample sizes ranging from 8 to 72, with similar proportions of male and female participants and average age. Treatment lasted from 4 weeks to 8 weeks. Table 2 presents the general information on the included trials.

**Table 2.**
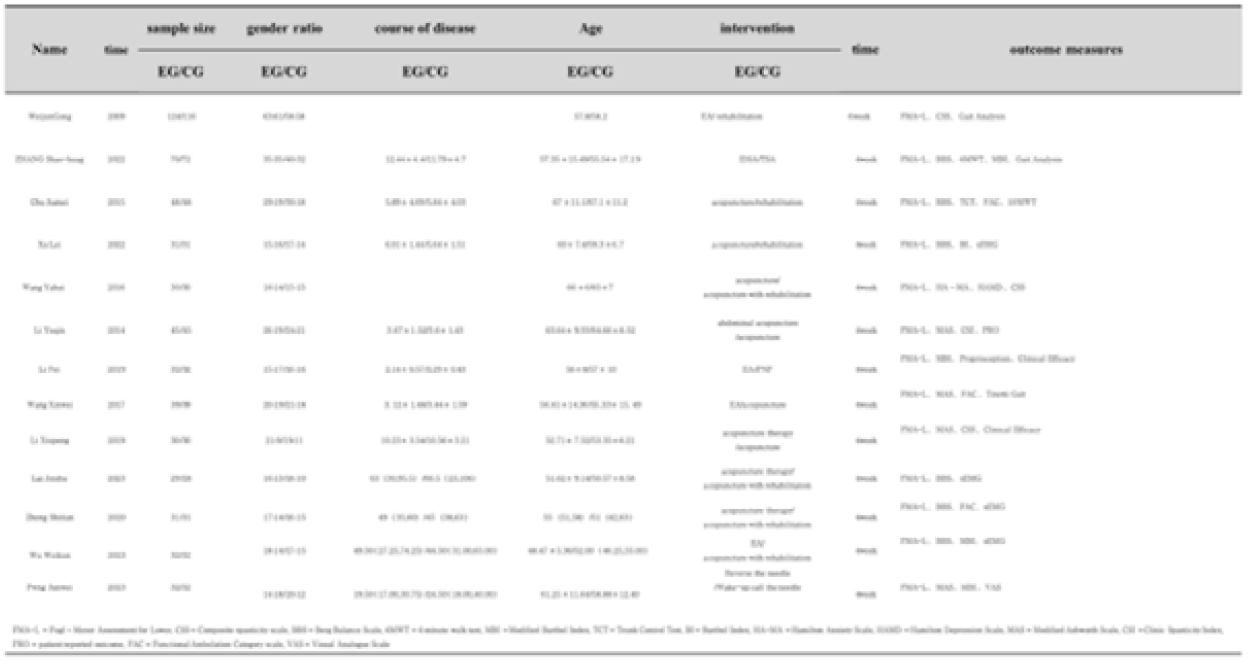
Basic characteristics of the included studies.

### 3.3 ROB in included studies

Cochrane Handbook was used to assess the literature included in this study. Twelve literary works were included, with ten trials^22-24,26-31^ (83.33%) having a low ROB in terms of the generation of random sequence, which was generated using a random number table or computer software. Two trials^25,33^ (16.67%) lacked detailed reports on the method used for random sequence generation, and one trials^28^ (8.33%) applied allocation concealment. Most trials lacked description for their blinding methods, but four^22,23,25,28^ (33.33%) reported blinding of outcome assessment. Assessment of selective reporting of results caused difficulty due to the absence of protocols for the included trials. Based on descriptions in their methodology section, all included trials had a low level of ROB. Thus, the overall quality of the 12 included trials was at a low risk (Figures 2A and 2B).

**Figure 1.**
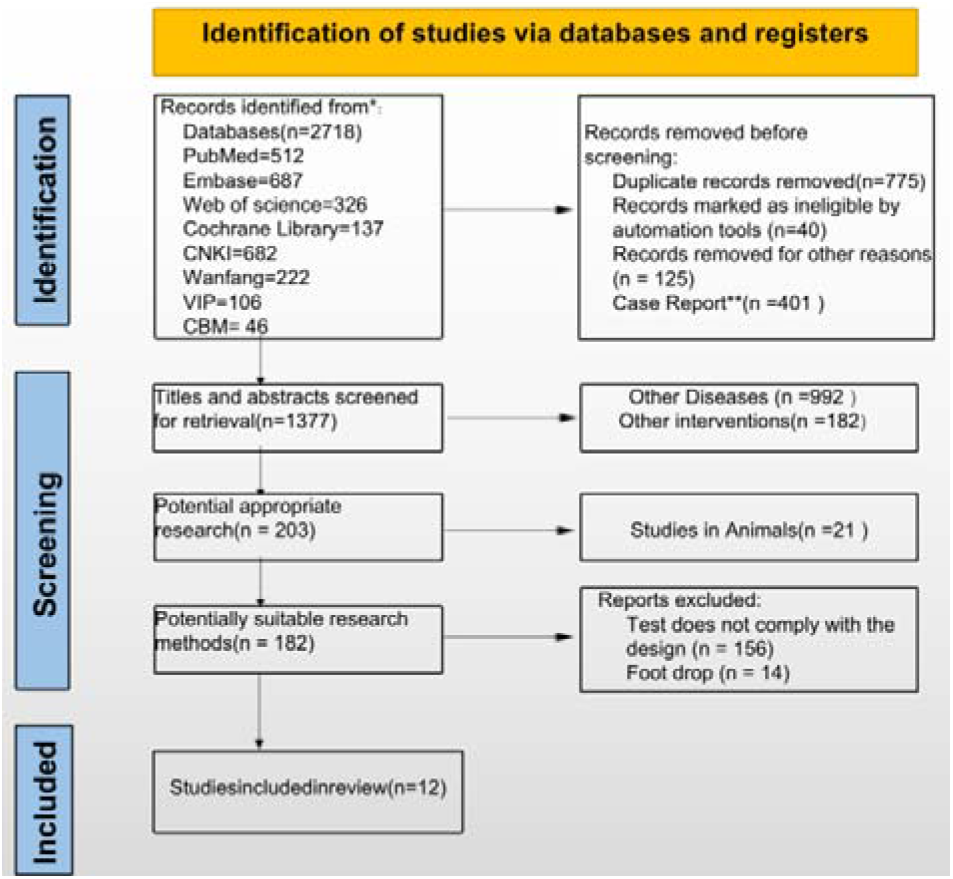
The flowchart of search results for meta-analysis.

**Figure 2.**
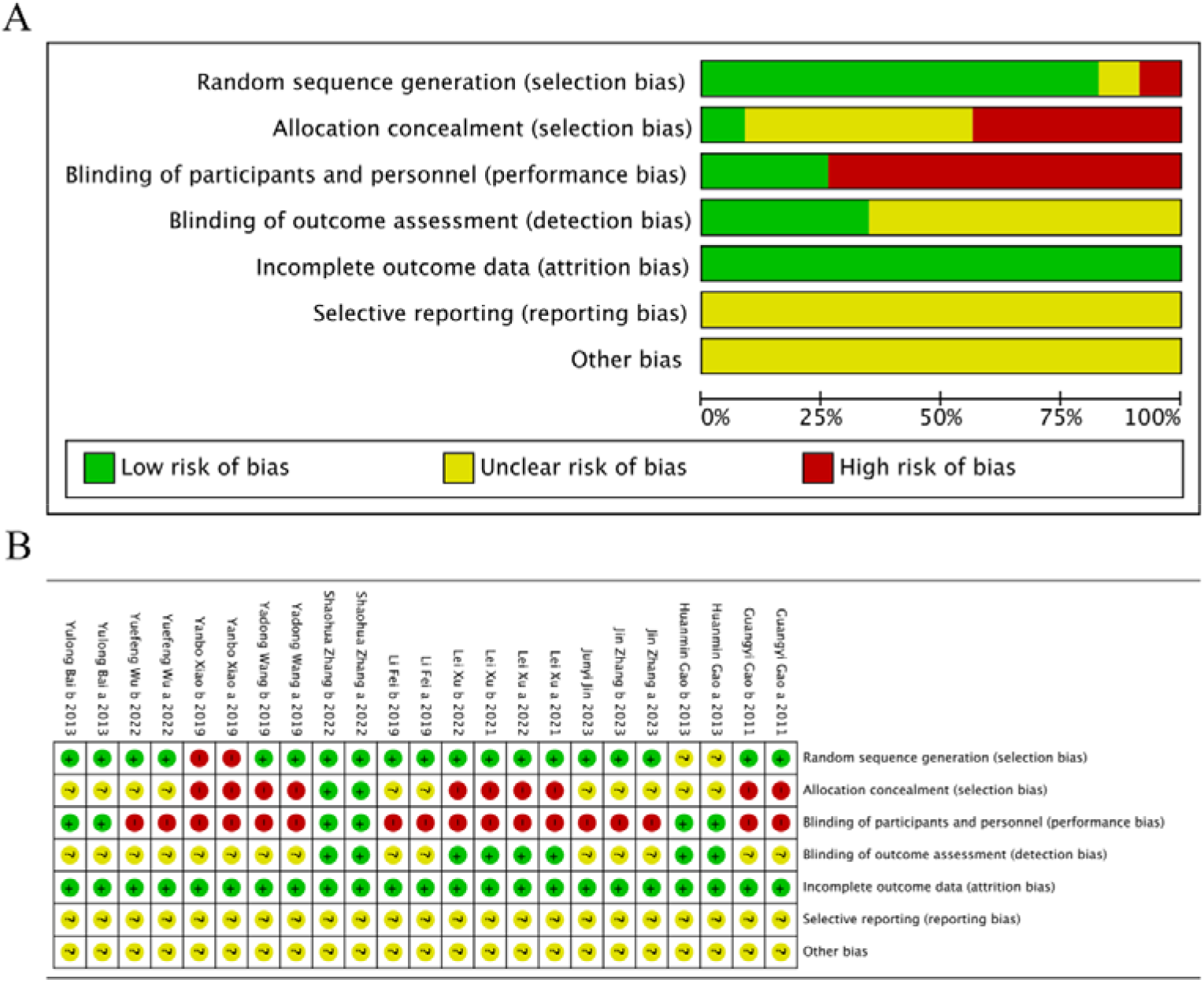
(A) Risk of bias graph. (B) Risk of bias summary.

### 3.4 Results Analysis

#### 3.4.1 FMA-L score

Eight trials^23,24,28-33^ (66.67%) involving 932 participants reported FMA-L as the primary outcome. Our findings showed a statistically significant the experimental group showed superior improvement in FMA-L compared with the control (SMD = −0.41, 95%CI[−0.84,-0.02],I^2^ = 89%,P < 0.05 random□effects mode)(Fig. 3A).There was significant heterogeneity. FMA-L included ≥10 studies, so meta□regression and subgroup analysis were used to explore the source of heterogeneity.

**Figure 3.**
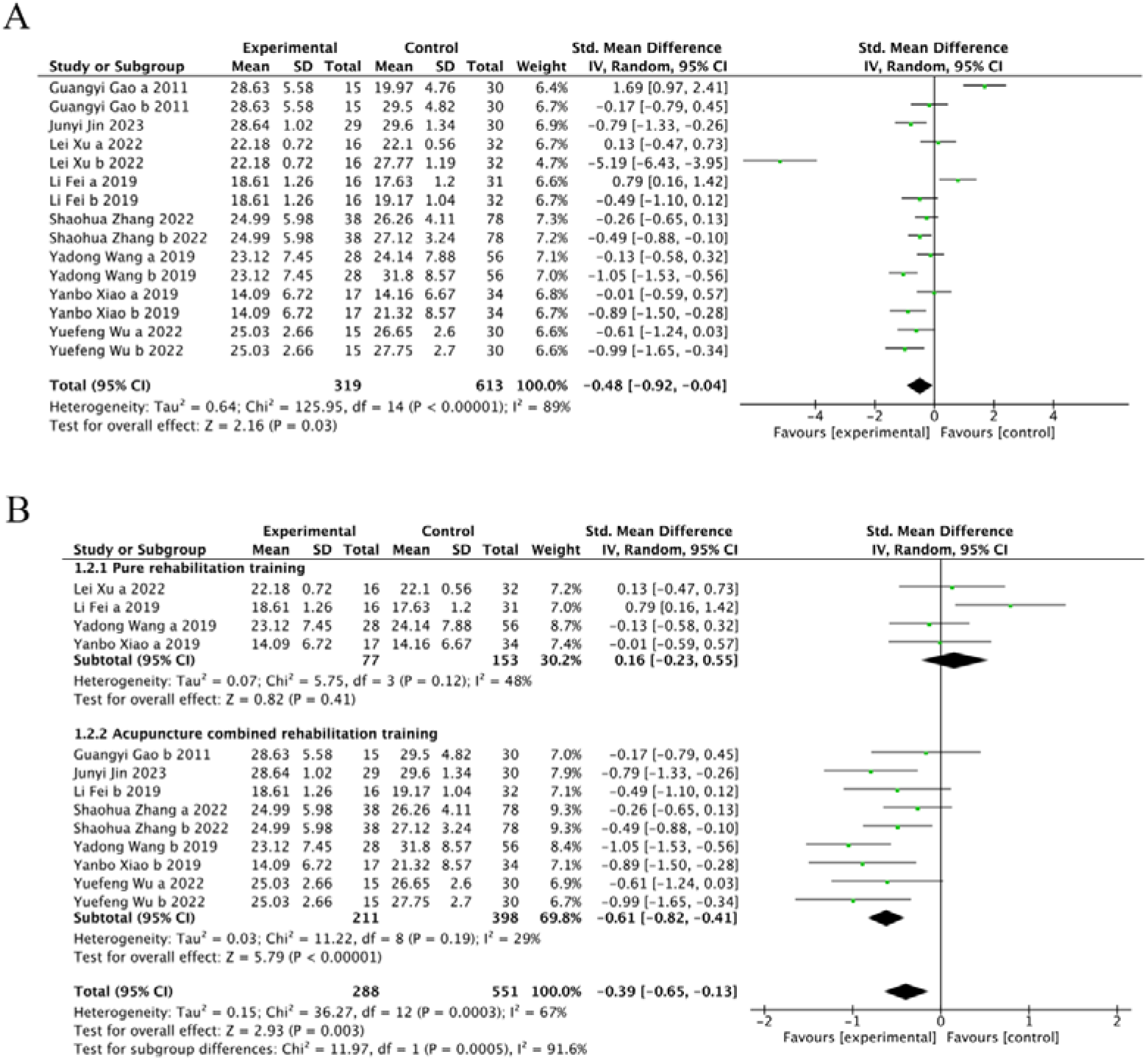
The forest plot of FMA-L. (A) Meta-analysis results containing FMA-L for all studies. (B) Results of subgroup analyses of FMA-L.

Meta□regression suggested that the tested variables including treatment time (P = 0.44), course of disease (P = 0.44), baseline level before treatment (p = 0.32) might not be the source of heterogeneity. Subgroup analyses were performed according to the different intervention modalities in the control group. The results of the subgroup analyses showed that there was no significant difference in the improvement of lower limb motor function between acupuncture and rehabilitation alone (SMD = 0.16, 95% CI [-0.23, 0.55], Z = 0.82, P = .41); however, acupuncture showed better improvement in lower limb motor function compared with combined treatment (SMD = −0.61, 95% CI [−0.82, −0.41], Z = 5.79, P < .001) (Fig. 3B). Sensitivity analysis revealed the results lacked robustness (see Fig. 4). When examining the possibility of publication bias, both the funnel plot (Fig. 5A) and Egger’s test (P = 0.368 > 0.05, Fig. 5B) indicated no evidence of such bias.

**Figure 4.**
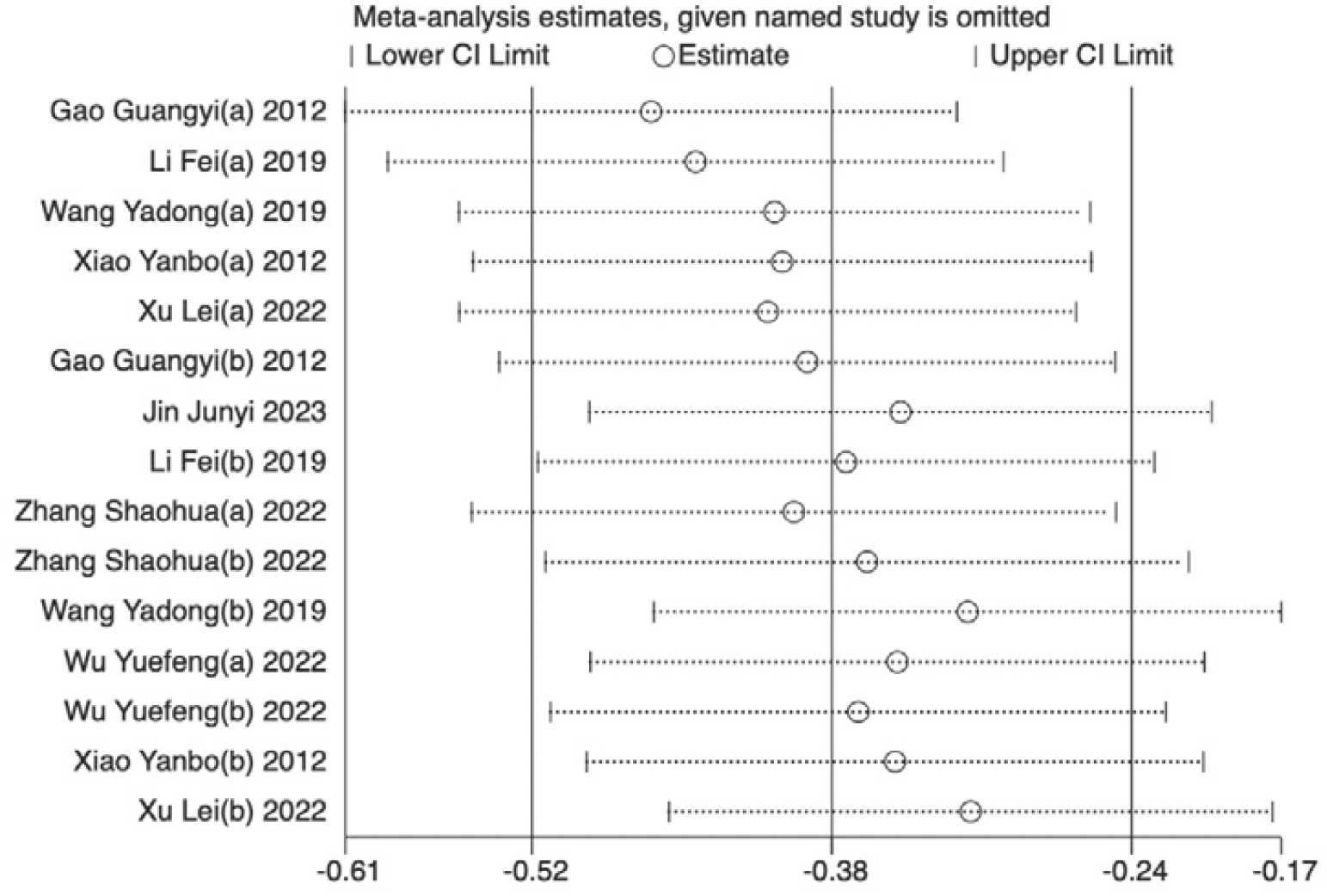
Sensitivity analysis results for FMA-L.

**Figure 5.**
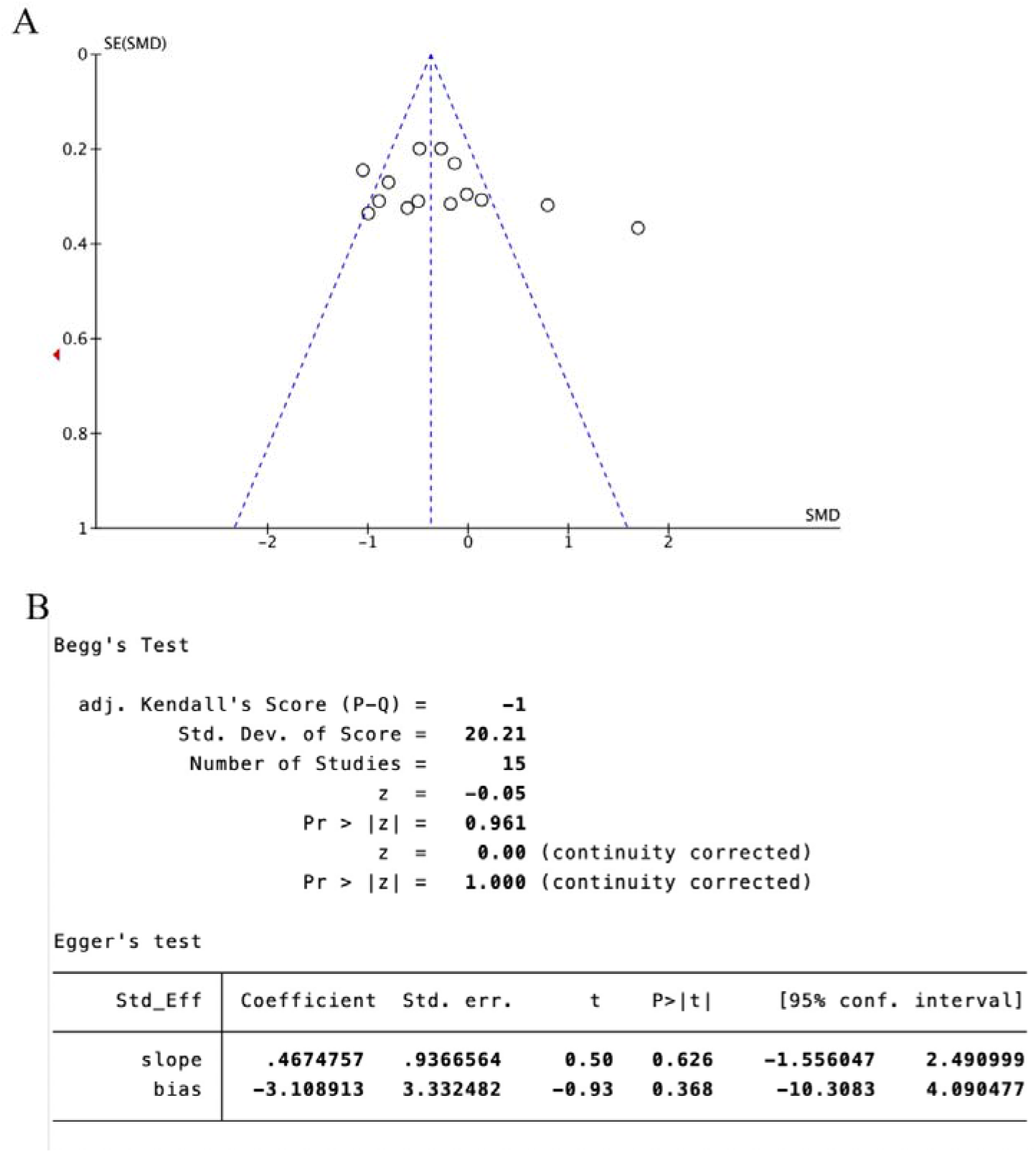
FMA-L scale publication bias test. (A) The funnel plot of FMA-L. (B) The Egger test result of FMA-L.

#### 3.4.2 BBS score

Four trials^22,23,27,28^ (33.33%) involving 570 participants reported BBS as the primary outcome. Our findings showed a statistically significant the experimental group showed superior improvement in BBS compared with the control (SMD = −0.85, 95%CI[−1.56,-0.07],I^2^ = 94%,P <.05, random□effects mode)(Fig. 6A).There was significant heterogeneity. BBS included ≥10 studies, so meta□regression and subgroup analysis were used to explore the source of heterogeneity. Meta□regression suggested that the tested variables including course of disease (P > .76), baseline level before treatment (P > .07) might not be the source of heterogeneity. However, we found that the control group (P > .03) may be the source of heterogeneity. Subgroup analyses were performed according to the different intervention modalities in the control group. The results of the subgroup analyses showed that acupuncture was a more effective intervention compared to rehabilitation alone in improving patients’ balance (SMD = 0.61, 95% CI [0.22, 1.00], Z = 3.09, P < 0.05); however, there was no statistically significant difference in the treatments when comparing with acupuncture combined with lower limb robotic exercise (SMD = −0.20, 95% CI [−0.47, 0.08], Z = 1.4, P = 0.16); whereas, we found that the acupuncture intervention was superior to acupuncture combined with rehabilitation exercises when compared to acupuncture combined with rehabilitation training. (SMD = −3.52, 95% CI [−4.19, −2.85], Z = 10.3, P < .001) (Fig. 6B). Sensitivity analysis revealed the results lacked robustness (see Fig. 7). When examining the possibility of publication bias, both the funnel plot (Fig.8 A) and Egger’s test (P = 0.382 > 0.05, Fig. 8B) indicated no evidence of such bias.

**Figure 6.**
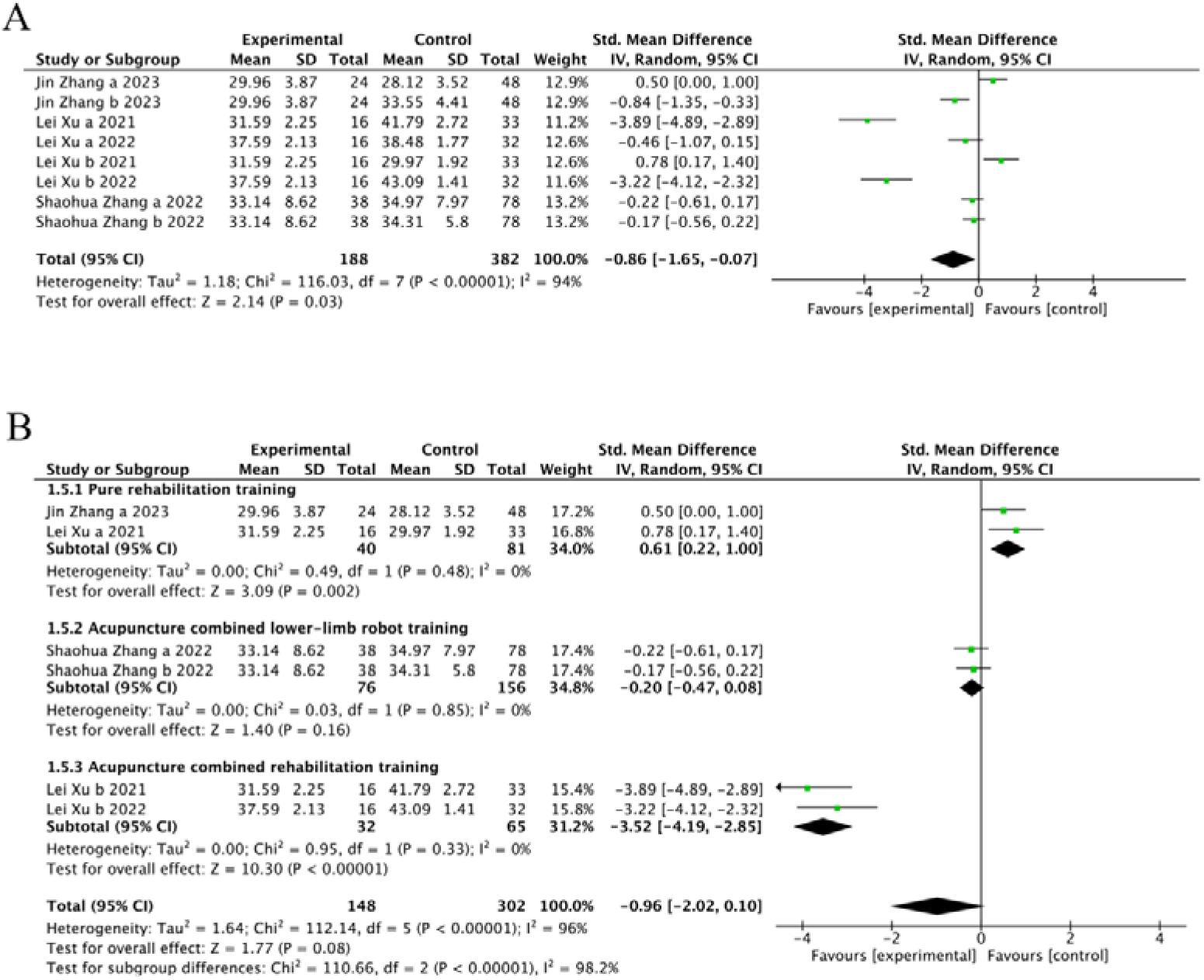
The forest plot of BBS. (A) Meta-analysis results containing BBS for all studies. (B) Results of subgroup analyses of BBS.

**Figure 7.**
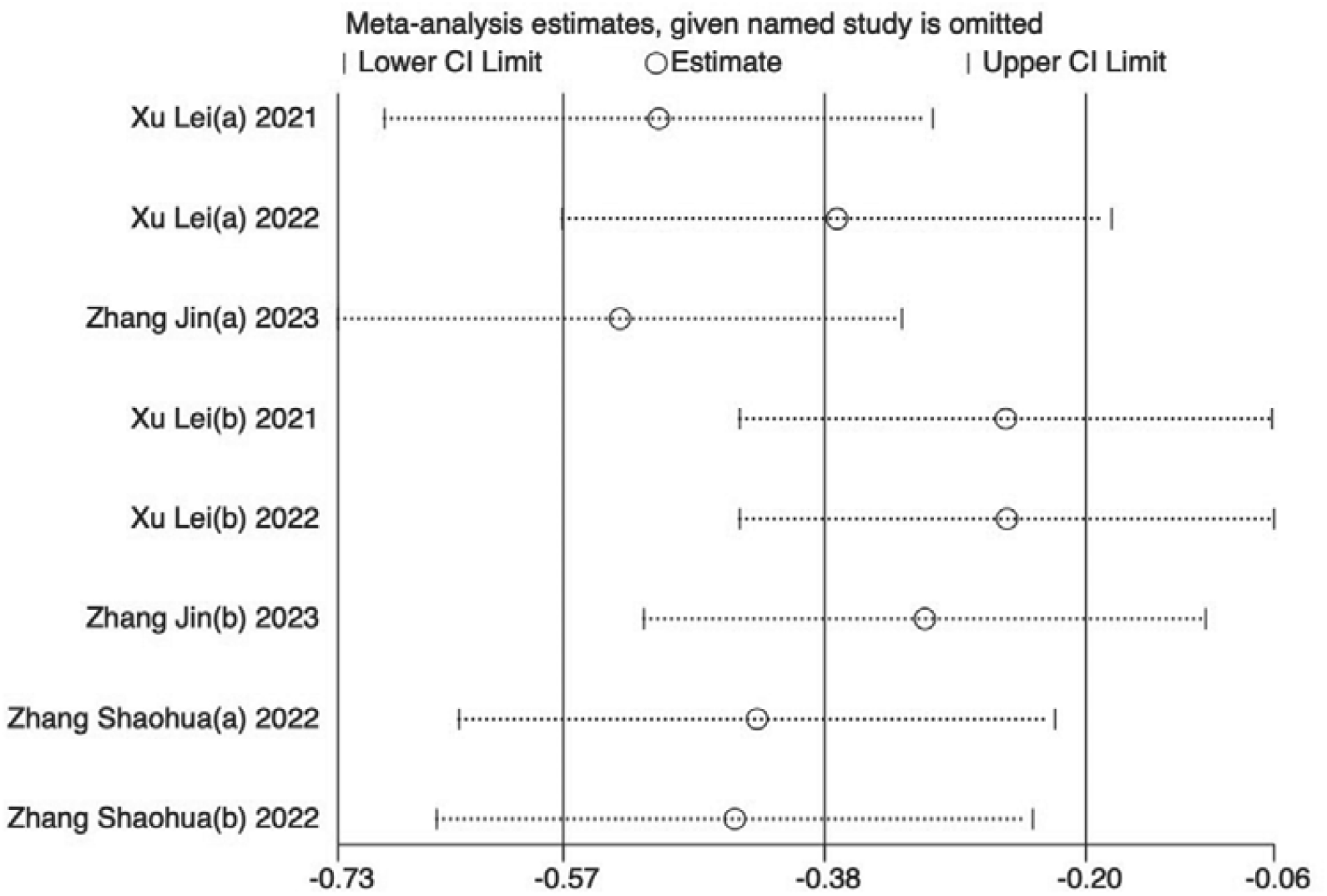
Sensitivity analysis results for BBS.

**Figure 8.**
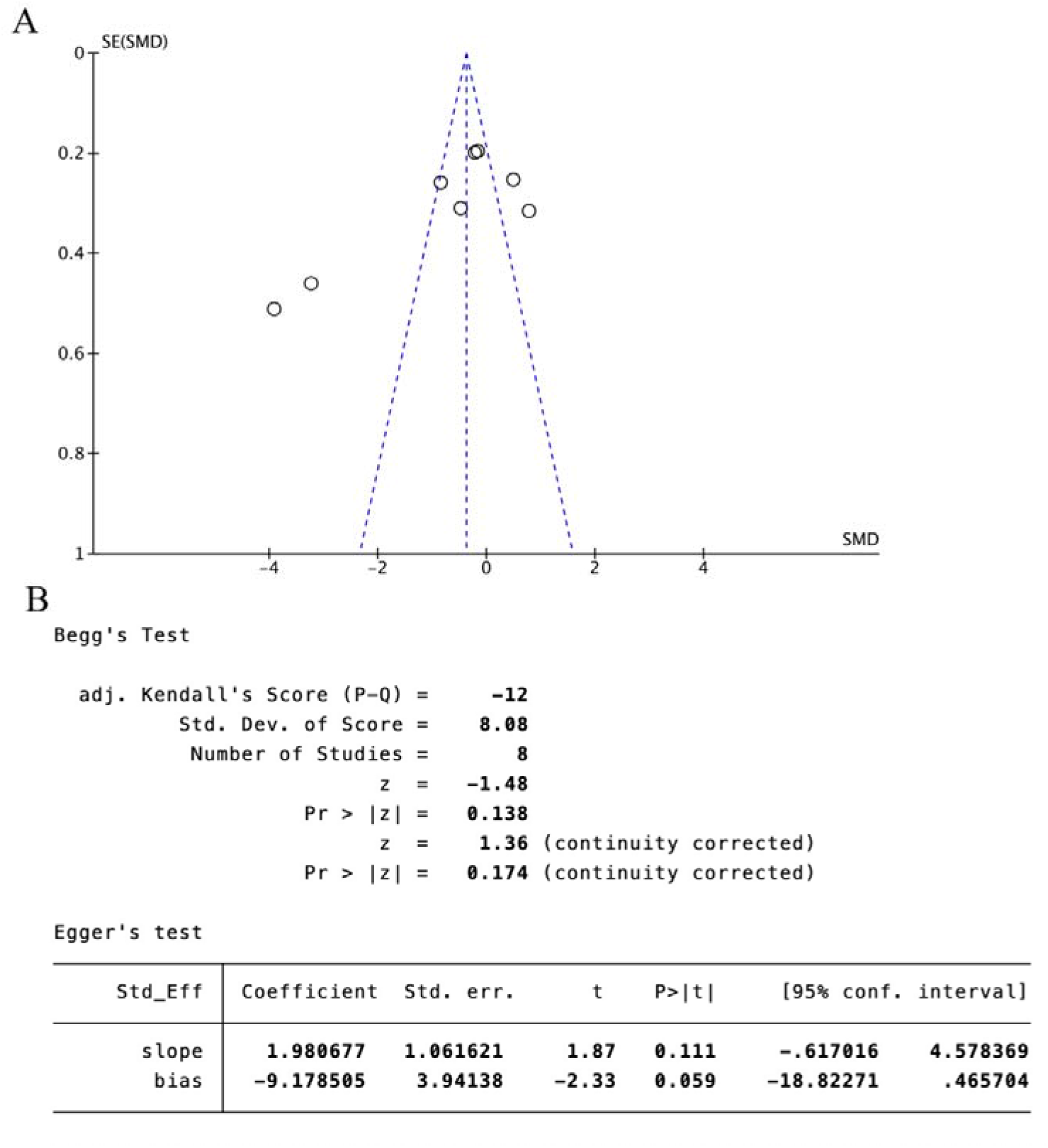
BBS scale publication bias test. (A) The funnel plot of BBS. (B) The Egger test result of BBS.

#### 3.4.3 MBI score

Five trials^24,25,28,30,32^ (41.67%) that involved 664 participants reported MBI as the primary outcome. Our findings showed a statistically significant the experimental group showed superior improvement in MBI compared with the control(SMD = 0.18, 95%CI[−0.40,-0.77],I^2^ = 91%,P = .54, random□effects mode) (Fig. 9A). There was significant heterogeneity. MBI included ≥10 studies, so meta□regression and subgroup analysis were used to explore the source of heterogeneity. Meta□regression suggested that the tested variables including course of disease (P > .39),course of disease (P > .14) might not be the source of heterogeneity. Subgroup analyses were performed according to the different intervention modalities in the control group. The results of subgroup analyses showed that acupuncture intervention was not statistically significant compared with rehabilitation alone (SMD = 0.2, 95% CI [−0.41, 0.8], Z = 0.63, P = .53), but acupuncture had a better therapeutic effect compared with combined rehabilitation training (SMD = −0.38, 95% CI [−0.6, −0.16], Z = 3.41, and P < .05) (Fig. 9B). Sensitivity analysis revealed the results lacked robustness (see Fig. 10). When examining the possibility of publication bias, both the funnel plot (Fig. 11A) and Egger’s test (P=0.018<0.05, Fig. 11B) indicated no evidence of such bias.

**Figure 9.**
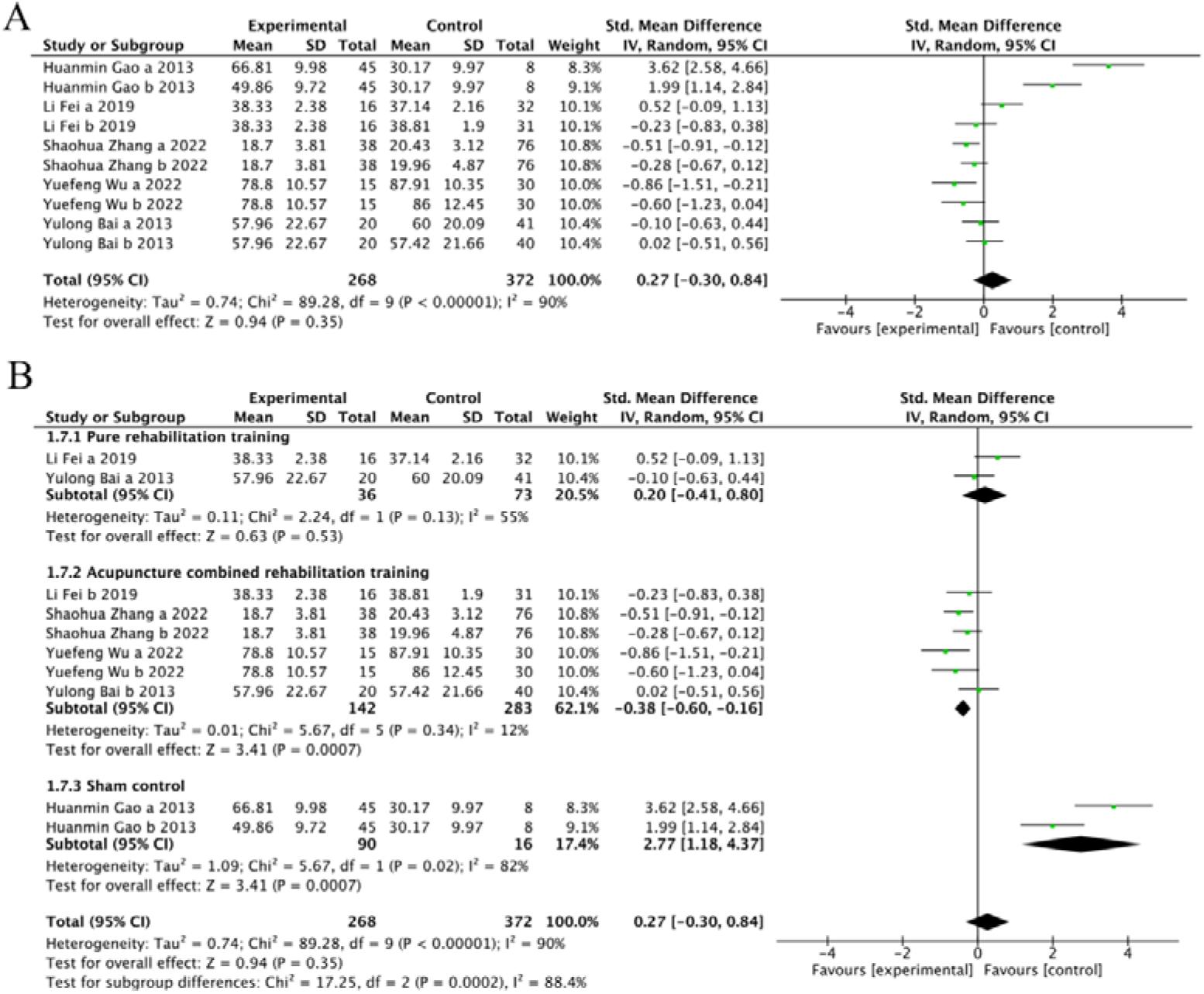
The forest plot of MBI. (A) Meta-analysis results containing MBI for all studies. (B) Results of subgroup analyses of MBI.

**Figure 10.**
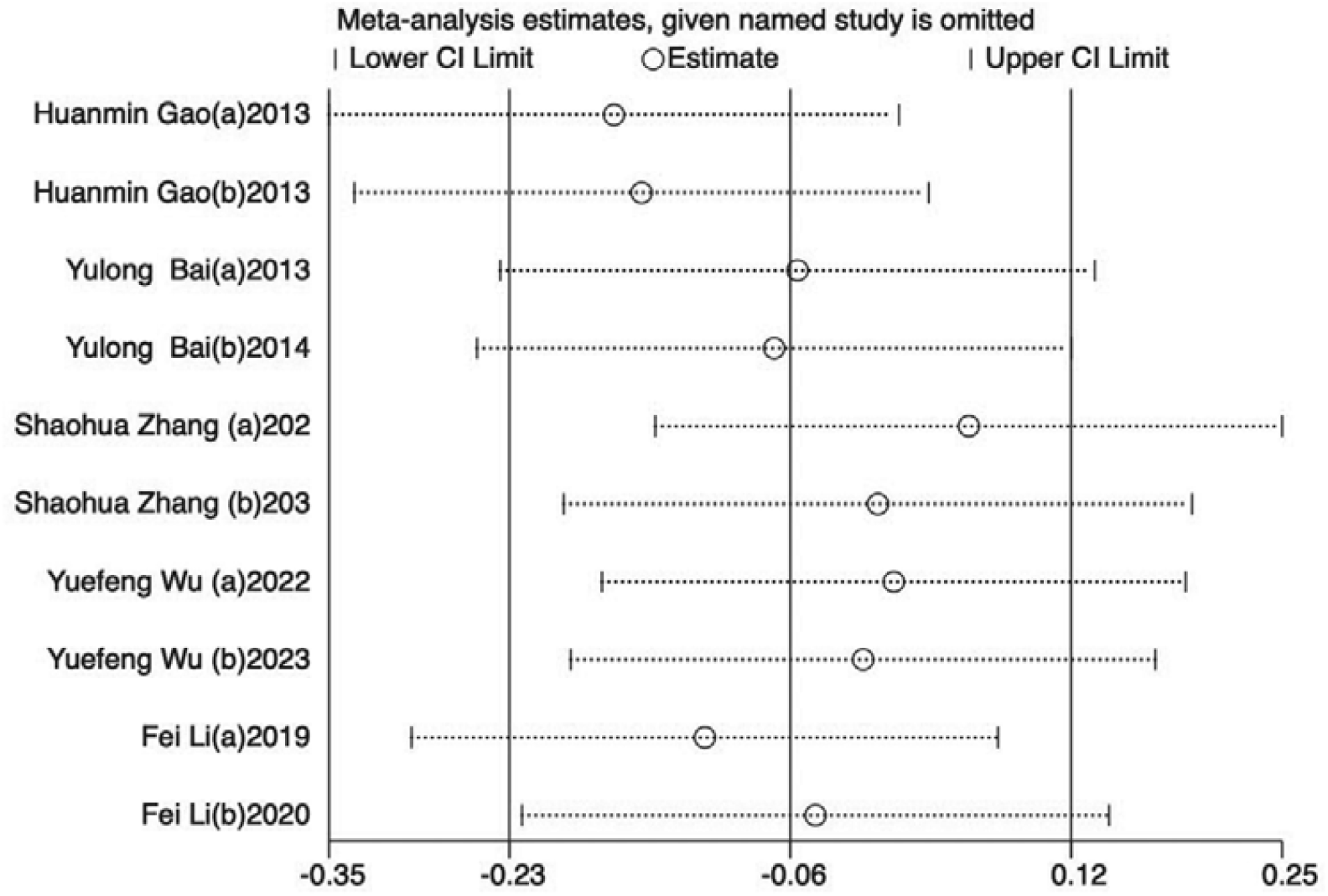
Sensitivity analysis results for MBI.

**Figure 11.**
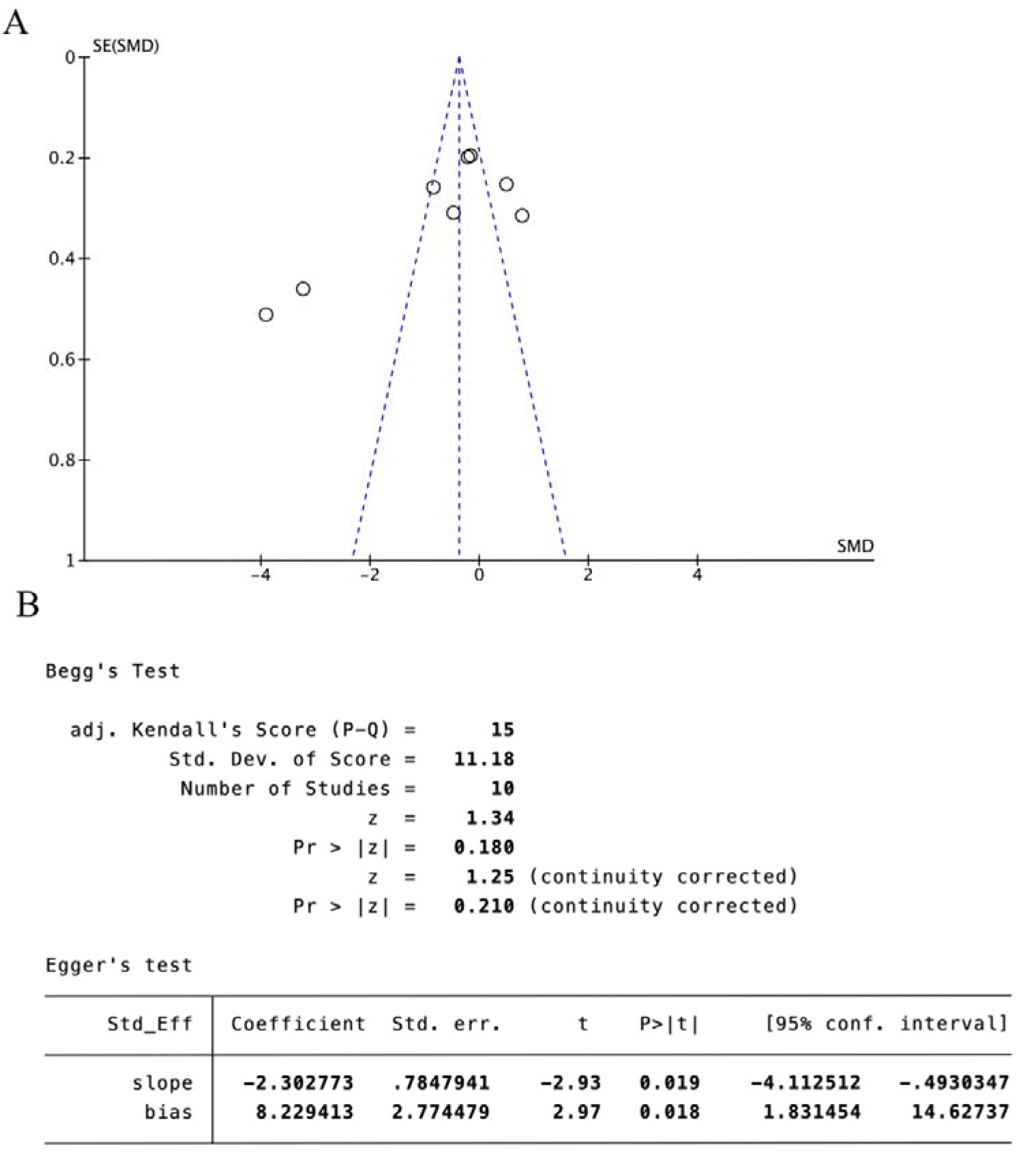
MBI scale publication bias test. (A) The funnel plot of MBI. (B) The Egger test result of MBI.

#### 3.44 FAC score

Two trials^27,31^ (16.67%) involving 312 participants reported FAC as the primary outcome. Our findings showed a not significant the experimental group showed superior improvement in FAC compared with the control(SMD=-0.74, 95%CI[−2.33,0.84],I^2^ = 97%,P = .36, random□effects mode) (Fig. 12A). There was significant heterogeneity. Subgroup analysis was used to explore the source of heterogeneity. Subgroup analyses were performed according to the different intervention modalities in the control group. The results of subgroup analyses showed that acupuncture intervention was not statistically significant compared with pure rehabilitation (SMD = 0.62, 95% CI [−0.96, 1.92], Z = 0.92, P = .35), and acupuncture had a not statistically significant therapeutic effect compared with combined rehabilitation training (SMD = −2.13, 95% CI [−4.58, 0.31], Z = 1.71, and P = .09) (Fig. 12B).

**Figure 12.**
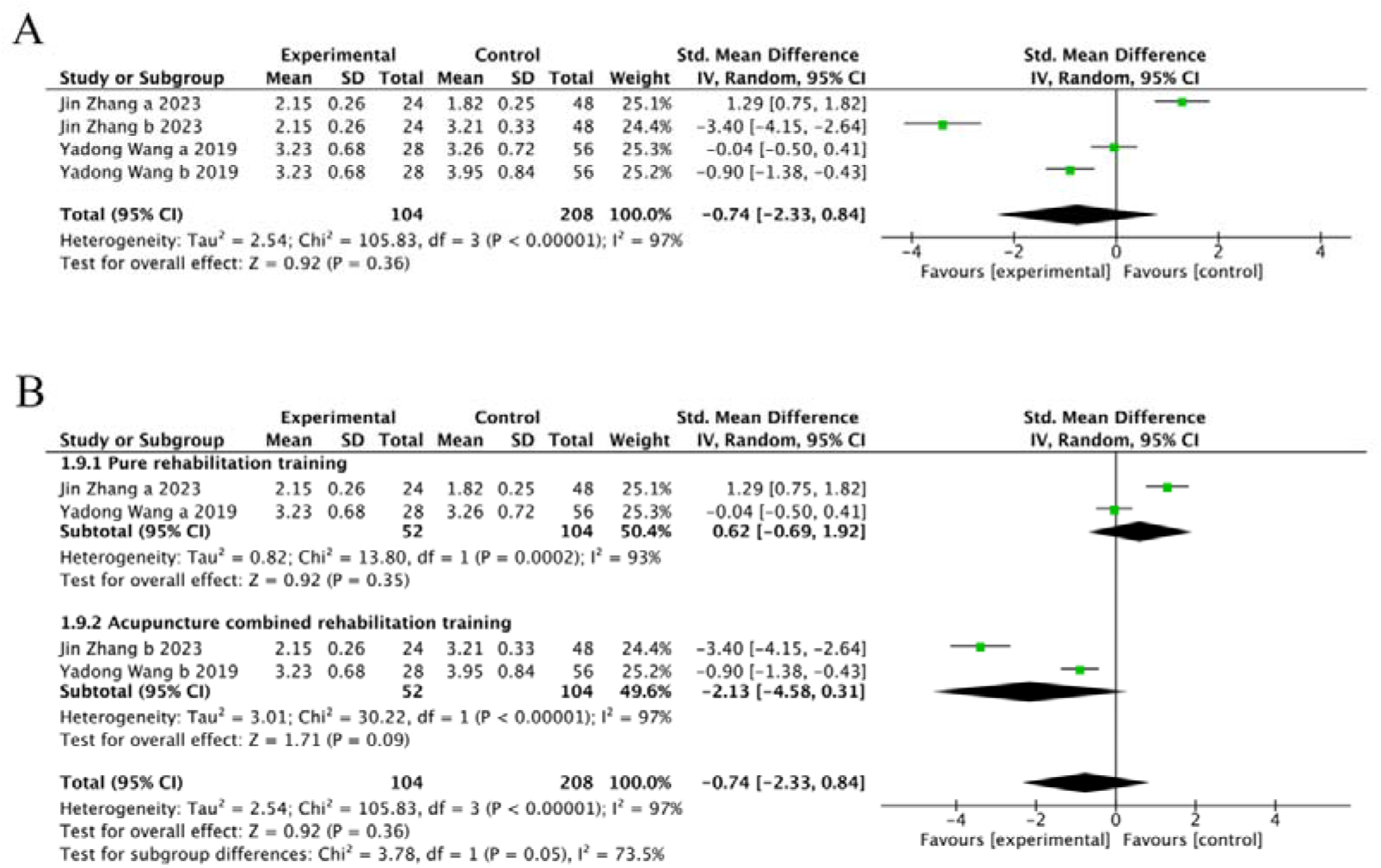
The forest plot of FAC. (A) Meta-analysis results containing FAC for all studies. (B) Results of subgroup analyses of FAC.

## 4 Discussion

In this review, we analyzed 12 trials that compared the effects of acupuncture or in combined rehabilitation training against pure rehabilitation training on LLMD. Our meta-analysis revealed that, compared to the control groups, acupuncture showed lesser efficacy in enhancing motor and balance functions. However, it was observed that acupuncture did not significantly impact the improvement of walking ability and daily living ability in patients with LLMD.

A meta-analysis was conducted on 8 articles that evaluated acupuncture treatment, using the FMA-L as the outcome measure. The literature included in this study had a wide range of control groups treatment intervention, with most trial interventions being pure rehabilitation training or acupuncture combined rehabilitation training. In the subgroup analysis based on the FMA-L scale, this variation served as the criteria for grouping. The findings indicated that the effectiveness of acupuncture treatment was comparable to that of solely rehabilitation training, with neither showing superiority over the combined approach of acupuncture and rehabilitation training. Furthermore, significant differences were observed among the subgroups, suggesting that the treatment intervention could contribute to the heterogeneity observed in this study. Sensitivity analysis, performed by sequentially excluding each study, confirmed the stability of these findings, despite the varied intervention approaches in the control groups.

Similarly, our subgroup analysis for BBS found that acupuncture was superior to pure rehabilitation training. Although at the overall evaluation, the experimental and control groups presented no difference in results. However, There were significant differences between different subgroups and reduced heterogeneity between groups. Therefore, the control group treatment interventions was one source of heterogeneity in this study. In addition, sensitivity analyses by the item-wise exclusion proved the robustness of these results.

In this study, there were five trials of MBI scales. Similarly, our subgroup analysis indicated that acupuncture was more effective than placebo control. Although the final results showed no apparent differences between the experimental and control groups, the differences between the different subgroups were statistically significant for the study. Therefore, the treatment intervention may be one source of heterogeneity in this study. The study also verifies the robustness of these results through sensitivity analyses conducted by item-wise exclusion.

Similarly, In the subgroup analysis of the FAC scale, control groups treatment intervention were used as the basis for grouping. Our subgroup analysis indicated that the experimental and control groups presented no difference in results when evaluated overall. In addition, heterogeneity between groups was reduced respectively. Therefore, the control group treatment intervention may be one source of heterogeneity in this study.Sensitivity analysis was conducted by excluding each literature one by one, indicating low sensitivity and robust meta-analysis results.

In the 12 randomized controlled trials analyzed, the treatment period ranged from 4 weeks to 12 weeks. Additionally, while 10 trials employed random assignment tables for participant allocation, only one trial implemented allocation concealment. And the lack of blinding of the participants and outcome assessor might result in bias in the outcomes. Despite these methodological shortcomings, a thorough bias analysis was conducted, leading to the conclusion that the overall risk of bias was low (p<0.05). Furthermore, sensitivity analyses and meta-analyses were performed on this body of literature, affirming the robustness of the results.

There was significant heterogeneity in the scale of FMA-L, BBS, FAC and MBI, respectively. First of all, the quality of the literature included in this study was universally imperfect due to many factors. The main factor is that the limitations of acupuncture treatment make it almost impossible to conduct RCT with the blinded experiment. Second, Subgroup analyses revealed that heterogeneity decreased with different intervention modalities in the control group, which may be due to differences in the rehabilitation training methods, acupuncture point selection, and stimulation volume that may interfere with clinical efficacy and assessment. Finally, The outcome measures such as the FMA-L, BBS, MBI, and FAC inherently depend on subjective judgment, which can be significantly influenced by the clinicians’ personal clinical experiences.

Previous literature has systematically evaluated motor function in hemiplegic patients after stroke, and in addition to methodological shortcomings, LLMD was only included as part of the study^34,35^. In our study, we conducted a comprehensive search employing stringent inclusion and exclusion criteria, along with recognized tools for assessing the quality of the included studies, to examine the efficacy of acupuncture in improving (LLMD) in post-stroke patients. Subsequently, a meta-analysis was performed to synthesize the findings. Despite the relatively low quality of some included studies, we systematically reviewed and analyzed the available evidence on the effectiveness of acupuncture for treating LLMD. Moreover, the study can provide support for the empirical evidence of acupuncture treatment in promoting the recovery of patients’ motor function in the future.

The limitations of the study mainly came from the control groups treatment intervention. The experimental group used pure acupuncture as the main intervention, but the specific type of acupuncture was not specified, and the complexity of the acupuncture treatment used in the included trials made it difficult to analyze subgroups based on the different types of acupuncture treatment. Moreover, The included literature does not restrict rehabilitation training to particular types. The pure rehabilitation training, placebo control, and acupuncture combined rehabilitation training mentioned in the article were also defined as control groups. However, further studies are necessary to confirm whether these treatments positively or negatively affect the efficacy of acupuncture. In addition, There was 1 studies comparing acupuncture versus placebo control, so the specific effect of acupuncture for LLMD was not clear.

At present, clinical trials of acupuncture for the treatment of LLMD still have many methodological limitations and the research results may be biased. In clinical practice, well-designed, large-sample, multi-center RCT are still required to further verify the effectiveness and benefits of acupuncture treatment. The study also aims to standardize the criteria for evaluating the clinical efficacy of scales such as FMA-L, BBS, and MBI, with attention to long-term outcomes. In addition, the optimal combination and timing of acupuncture combined with rehabilitation training needs to be further explored to provide support to the recovery of patients’ motor function.

## 5 Conclution

This study proved that the efficacy of acupuncture for LLMD is inconclusive. But, the results of this study suggest that acupuncture combined rehabilitation training has clear superiority for LLMD compared with pure acupuncture or pure rehabilitation training, for provide evidence-based support for further research and promote the effects of acupuncture treatment on LLMD in poststroke patients. Influenced by the quality and quantity of the included literature, future studies should focus on better-designed and more comprehensive outcome indicators as well as multi-center and high-quality RCT.

## Data Availability

All data generated or analyzed during this study are included in this published article (and its supplementary information files.

## Author contributions

All authors contributed to the study conception and design

Data curation: Peng Chen and Xing Jin

Formal analysis: Peng Cheng, Xing Jig and Debiao Yu

Funding acquisition: Bin Shao Investigation: Yu Debiao and Yaoyu Lin

Methodology: Bin Shao, Debiao Yu, and Xiaotin Chen

Resources: Xing Jin, Debiao Yu, and Peng Cheng

Software: Xiaotin Chen, and Yaoyu Lin

Validation: Bin Sha and Debiao Y

Visualization: Debiao Y and Xiaotin Chen

Writing – original draft: Peng Cheng and Debiao Y

Writing – review & editing: Xing Jin and Debiao Yu

## Abbreviations

LLMD: lower limb motor dysfunction
95%CI: 95% confidence interval
FMA-L: Fugl–Meyer Assessment for Lower
BBS: Berg Balance Scale
FAC: functional ambulation category scale
MBI: Modified Barthel Index
OR: odds ratio
RCT: randomized controlled trial
REM: random-effects model
SMD: standardized mean difference

## References

1. Barthels D, Das H. Current advances in ischemic stroke research and therapies. Biochim Biophys Acta Mol Basis Dis. Apr 1 2020;1866(4):165260. doi:10.1016/j.bbadis.2018.09.012

2. Global, regional, and national burden of stroke and its risk factors, 1990-2019: a systematic analysis for the Global Burden of Disease Study 2019. Lancet Neurol. Oct 2021;20(10):795–820. doi:10.1016/s1474-4422(21)00252-0

3. Katan M, Luft A. Global Burden of Stroke. Semin Neurol. Apr 2018;38(2):208–211. doi:10.1055/s-0038-1649503

4. Veldema J, Gharabaghi A. Non-invasive brain stimulation for improving gait, balance, and lower limbs motor function in stroke. J Neuroeng Rehabil. Aug 3 2022;19(1):84. doi:10.1186/s12984-022-01062-y

5. Gibbs ME, Richdale AL, Ng KT. Biochemical aspects of protein synthesis inhibition by cycloheximide in one or both hemispheres of the chick brain. Pharmacol Biochem Behav. Jun 1979;10(6):929–31. doi:10.1016/0091-3057(79)90069-8

6. Grau-Pellicer M, Chamarro-Lusar A, Medina-Casanovas J, Serdà Ferrer BC. Walking speed as a predictor of community mobility and quality of life after stroke. Top Stroke Rehabil. Jul 2019;26(5):349–358. doi:10.1080/10749357.2019.1605751

7. Yoo YJ, Lim SH. Assessment of Lower Limb Motor Function, Ambulation, and Balance After Stroke. Brain Neurorehabil. Jul 2022;15(2):e17. doi:10.12786/bn.2022.15.e17

8. Knutson JS, Fu MJ, Sheffler LR, Chae J. Neuromuscular Electrical Stimulation for Motor Restoration in Hemiplegia. Phys Med Rehabil Clin N Am. Nov 2015;26(4):729–45. doi:10.1016/j.pmr.2015.06.002

9. Kwakkel G, Veerbeek JM, van Wegen EE, Wolf SL. Constraint-induced movement therapy after stroke. Lancet Neurol. Feb 2015;14(2):224–34. doi:10.1016/s1474-4422(14)70160-7

10. Oh ZH, Liu CH, Hsu CW, Liou TH, Escorpizo R, Chen HC. Mirror therapy combined with neuromuscular electrical stimulation for poststroke lower extremity motor function recovery: a systematic review and meta-analysis. Sci Rep. Nov 16 2023;13(1):20018. doi:10.1038/s41598-023-47272-9

11. Birch S, Robinson N. Acupuncture as a post-stroke treatment option: A narrative review of clinical guideline recommendations. Phytomedicine. Sep 2022;104:154297. doi:10.1016/j.phymed.2022.154297

12. Qin S, Zhang Z, Zhao Y, et al. The impact of acupuncture on neuroplasticity after ischemic stroke: a literature review and perspectives. Front Cell Neurosci. 2022;16:817732. doi:10.3389/fncel.2022.817732

13. Zhang J, Lu C, Wu X, Nie D, Yu H. Neuroplasticity of Acupuncture for Stroke: An Evidence-Based Review of MRI. Neural Plast. 2021;2021:2662585. doi:10.1155/2021/2662585

14. Chavez LM, Huang SS, MacDonald I, Lin JG, Lee YC, Chen YH. Mechanisms of Acupuncture Therapy in Ischemic Stroke Rehabilitation: A Literature Review of Basic Studies. Int J Mol Sci. Oct 28 2017;18(11)doi:10.3390/ijms18112270

15. Zhu ZL, Shen TY, Li XX, Mao JF, Xie T, Zhang JB. [Progress of researches on involvement of corticospinal tract in the effect of acupuncture on improvement of post-stroke motor dysfunction]. Zhen Ci Yan Jiu. Sep 25 2022;47(9):843–6. doi:10.13702/j.1000-0607.20210670

16. Liu J, Zhao G, Niu Y, Gan T, Yan Z, Zhang Y. Effect of electro-acupuncture therapy on limb spasm and excitability of motor neurons in stroke rats. Zhejiang Da Xue Xue Bao Yi Xue Ban. Jun 25 2021;50(3):361–368. doi:10.3724/zdxbyxb-2021-0007

17. Shen Y, Hu L, Ge J, Li L. Effect of electroacupuncture treatment combined with rehabilitation care on serum sirt3 level and motor function in elderly patients with stroke hemiparesis. Medicine (Baltimore). Apr 14 2023;102(15):e33403. doi:10.1097/md.0000000000033403

18. Zeng You-hua BY-h, GE Fang. Clinical observation on eye acupuncture combined with bodyacupuncture therapy treating lower limb spasticity after stroke. China Journal of Traditional Chinese Medicine and Pharmacy. 2021;36(01):558-560(in chinese).

19. Zhu Y, Zhang L, Ouyang G, et al. Acupuncture in subacute stroke: no benefits detected. Phys Ther. Nov 2013;93(11):1447–55. doi:10.2522/ptj.20110138

20. Gladstone DJ, Danells CJ, Black SE. The fugl-meyer assessment of motor recovery after stroke: a critical review of its measurement properties. Neurorehabil Neural Repair. Sep 2002;16(3):232–40. doi:10.1177/154596802401105171

21. Downs S. The Berg Balance Scale. J Physiother. Jan 2015;61(1):46. doi:10.1016/j.jphys.2014.10.002

22. Xu L, Li F, Wang M, Yan XZ, D. Yh. [Scalp acupuncture combined with suspension training for balance dysfunction in patients with stroke: a randomized controlled trial]. Zhongguo Zhen Jiu. Dec 12 2021;41(12):1308–12. doi:10.13703/j.0255-2930.20201217-0002

23. Xu L, Li L, Du JT, Tong X, Lu Y, Li F. [Effect of acupuncture at Huatuo Jiaji (EX-B2) combined with core muscle training on motor function of lower limbs in patients with hemiplegia after stroke]. Zhen Ci Yan Jiu. Feb 25 2022;47(2):154–9. doi:10.13702/j.1000-0607.20210487

24. Li F, Sun Q, Shao XM, et al. [Electroacupuncture combined with PNF on proprioception and motor function of lower limbs in stroke patients: a randomized controlled trial]. Zhongguo Zhen Jiu. Oct 12 2019;39(10):1034–40. doi:10.13703/j.0255-2930.2019.10.002

25. Gao H, Li X, Gao X, Ma B. Contralateral needling at unblocked collaterals for hemiplegia following acute ischemic stroke. Neural Regen Res. Nov 5 2013;8(31):2914–22. doi:10.3969/j.issn.1673-5374.2013.31.004

26. Bai YL, Li L, Hu YS, et al. Prospective, randomized controlled trial of physiotherapy and acupuncture on motor function and daily activities in patients with ischemic stroke. J Altern Complement Med. Aug 2013;19(8):684–9. doi:10.1089/acm.2012.0578

27. Zhang Jin CM, KAN Wen. Effects of acupuncture-moxibustion combined with motorimagery therapy on surface electromyography and center ofmass trajectory during walking in patients with cerebralinfarction in recovery stage. Shanghai Journal of Acupuncture and Moxibustion. 2023;42(01):6-11(in chinese).

28. Zhang SH, Wang YL, Zhang CX, et al. Effects of Interactive Dynamic Scalp Acupuncture on Motor Function and Gait of Lower Limbs after Stroke: A Multicenter, Randomized, Controlled Clinical Trial. Chin J Integr Med. Jun 2022;28(6):483–491. doi:10.1007/s11655-021-3525-0

29. Junyi J. Clincal observation of double acupuncture combined with exercise therapy in the teratment of lower limb dyskonesia in the convalescent stage of stroke. 2022;

30. Wu Yuefeng CL, DONG Xiaogiong, ZHU Tong, GAO Haijun, ZHANG Fang, XU Xiabin, TAO Ligin. Scalp acupuncture combined with robot-assisted walkingtraining improves the gait of stroke patients. Zhejiang Medical Journal. 2022;44(03):269-273+278(in chinese).

31. Wang Yadong YD. The Effect of Acupuncture and Moxibustion Combined with Rehabilitation Training on Functional Recovery of the Lower Limbs in Hemiplegic Stroke Patients. Neural Injury and Functional Reconstruction. 2019;14(02):102-103(in chinese).

32. Xiao Yanbo TY, MA Haibin. Acupuncture and moxibustion combined with rehabilitation training in the treatment of stroke 102 cases of efficacy observation. Ningxia Medical Journal. 2012;34(06):561-562(in chinese).

33. Guanyi G. Clinical Research of Scalp Acupuncture & Facilitation TechniquesTherapy on Hemiplegic Patients with ischemic Stroke. 2011;

34. Wang Y, Lu M, Liu R, et al. Acupuncture Alters Brain’s Dynamic Functional Network Connectivity in Stroke Patients with Motor Dysfunction: A Randomised Controlled Neuroimaging Trial. Neural Plast. 2023;2023:8510213. doi:10.1155/2023/8510213

35. Liu Y, Tang Y, Wang L, et al. Optimal acupuncture methods for lower limb motor dysfunction after stroke: a systematic review and network meta-analysis. Front Neurol. 2024;15:1415792. doi:10.3389/fneur.2024.1415792

